# Effectiveness of single-dose use of oral cholera vaccine towards reducing cholera incidence and severity during 2022-2023 in Malawi: A cross-sectional study

**DOI:** 10.64898/2026.03.24.26349236

**Authors:** Selemani Ngwira, Alex Thawani, Vincent Kamfozi, Dzinkambani Kambalame, Randy Mungwira, Flora Dimba, Mike Chisema, Gertrude Chapotera, Ernest Ulaya, Annie Mwale, Isaach Ndemera, Joseph Wu, Wiseman Chimwanza, Matthew Kagoli, Vincent Samuel Phiri

**Affiliations:** Luke International; Public Health Institute of Malawi; World Health Organisation; Kamuzu University of Health Science; Expanded Program on Immunisation -Ministry of Health; Presidential Task Force on Public Health Emergencies; Birkbeck, University of London

**Keywords:** Oral Cholera Vaccination, single dose, cholera, outbreak, vaccine effectiveness

## Abstract

**Background:** Approximately 59,000 cases and 1700 deaths were reported during the 2022-2023 cholera outbreak in Malawi. In response, the Ministry of Health implemented Oral Cholera vaccination campaigns (OCV) as one of the interventions. Four series of single-dose reactive OCV campaigns were conducted in 21 health districts between May 2022 and September 2023. OCV survey report of 2023 estimated a coverage of 55.40%. It was barely known how a single dose of OCV interrupts community transmission. This study was conducted to provide evidence on the use of a single-dose strategy and its contribution towards reducing the risk of cholera infection.

**Method:** A cross-sectional quantitative analysis was conducted to assess the effectiveness of OCV in reducing cholera incidence, severity, and mortality during the 2022-2023 cholera outbreak in Malawi. A national cholera line-list was used for analysis.

**Results:** Oral cholera vaccination coverage was at 2.0% of 28,920 suspected cholera cases. The effectiveness of a single dose of OCV towards reducing cholera infections was 98.00%. and associated with lower odds of severe dehydration (OR = 0.50; 95% CI: 0.39–0.64), with OCV effectiveness at 50%(95% CI: 36-61) Case fatality rate among vaccinated was 1.20 (95% CI: 0.54; 2.6, p = 0.025) and among unvaccinated it was 2.80% (95% CI: 2.6; 3.0, p = 0.025).

**Conclusion:** A single dose of the OCV campaign conducted during the 2022-2023 cholera outbreak in Malawi contributed to the reduction of cholera incidence. The intervention complemented other long-term interventions such as Water, Sanitation and Hygiene Oral Dehydration Points, Case Area Targeted Intervention, and Risk Communication and Community Engagements.

## Introduction

Cholera outbreaks remain a public health problem, especially in limited-resource countries. Currently, the global community is responding to the seventh cholera pandemic. Every year, approximately 1 million individuals are at risk of cholera[1]. Globally, 518,328 cases and 6508 deaths were reported between January 2025 and 28 Sept 2025, of which 199,914 cases were reported from 21 countries, and 4,481 deaths from 18 countries in Africa[2].

The actual burden of cholera disease in Africa is barely known. Delay in detection of cholera outbreaks and underreporting of cases and deaths contribute to low estimates of the disease burden. This problem persists due to poor surveillance systems in most African countries[3].

Populations with limited access to water, Residents in slums, refugee camps, and areas with open defecation are at high risk and prone to cholera outbreaks[4]. During cholera outbreaks, cholera transmission is exacerbated by multiple factors, including being in contact with a confirmed case, being male, intermittent availability of safe water, and high density of pit latrines[5]. Cholera outbreak in these settings is inevitably due to the presence of the 0139 Sero-group in circulation in the environment[6].

Cholera is endemic in Malawi. The first cholera outbreak was reported in 1973. Since then, the country has experienced recurring yearly cholera outbreaks[7,8]. The highest numbers of cholera cases and deaths were reported in 2022-2023, with over 59,000 cases and 1700 deaths, followed by 2001-2002, with 33,564 cases and 968 deaths[7]. The main prevalent serogroup for the 2022 -2023 cholera outbreak was Vibrio cholerae 01-strain Ogawa[9].

The Malawi government, through the Ministry of Health, with support from partners, implemented multiple interventions during the 2022-2023 cholera outbreak. Such intervention include scaling up capacity building on cholera case management, surveillance, Infection Prevention and Control, across all levels, intensified Risk Communication and Community Engagement(RCCE) activities such as, visual aids, radio programs, airing of informative jingles, engaging local leaders and religious figures, and van-publicity; upgrading Cholera Treatment Centres or Units(CTC/CTU), setting up of Oral Rehydration Points(ORP), rehabilitation and drilling new boreholes, conducting Case Area Targeted Interventions(CATI) and Oral Cholera Vaccination Campaigns(OCV)[10].

There are three types of Oral Cholera Vaccine prequalified by WHO: Dukoral, Euvichol-plus, and Euvichol-S. Euvichol-plus and Euvichol-S are suitable for over one year and above. The two vaccines are commonly used for mass vaccination and in reactive campaigns during cholera outbreaks[11].

During the 2022-2023 Cholera outbreak, the Malawi government used Euvichol-plus for a reactive campaign targeting 21 districts. The campaign employed a single-dose strategy due to the global shortage of OCV[11,12]. Euvichole-plus has shown effectiveness in reducing cholera incidence in cholera outbreaks setting. In Bangladesh, the effectiveness of two doses of Euvichol-plus was estimated at 66% (99.5% CI: 30 to 83) against medically attended cholera cases[13]. While in DRC using a single dose strategy, the effectiveness of Euvichol-plus was estimated at 52.7% (95% CI: 31 to 67.4) for 12 to 17 months[14]. The effectiveness of the 2022-2023 reactive campaign using a single dose of Euvichol-plus in Malawi is barely documented. To fill in this gap of knowledge, this study assessed the effectiveness of using a single dose of oral cholera vaccine towards reducing cholera incidence during the 2022-2023 cholera outbreak in Malawi.

## Materials and Methods

### study design and population

Quantitative cross-sectional analytical methods were employed to estimate the effectiveness of the OCV in reducing both the incidence and severity of cholera, based on data from the national cholera line-list during the 2022-2023 cholera outbreak. The Ministry of Health reported 59,088 cholera cases, both suspected and laboratory-confirmed cases[10,15]. All these cholera cases were captured at health facilities using a cholera line list and register book. A national cholera line list was our sampling frame. It included both suspected and confirmed cholera cases that were documented in all 29 health districts. The records were accessed on 25^th^ January 2025

### Sample Size

The study analysed 28,920 records of cholera cases from the national cholera line list with known vaccination and disease outcome status.

### Data Analysis

R software™ version 4.4.3 was employed for data cleaning, analysis and visualisation. Analysis included observations with known vaccination status and disease outcome. Descriptive statistics were calculated for the frequency and proportion of vaccinated individuals among cholera cases. Variables of interest were disease outcome, age, dehydration level, sex, occupation, district, district population and vaccination status. A chi-square test was used to assess the significance of the association among variables. Multivariable analysis was conducted using logistic regression to estimate odds ratios. The 2022 population from the National Statistical Office was used to calculate the Attack Rate (AR) among vaccinated cases and unvaccinated cases. Vaccine effectiveness (VE) in reducing cholera infection was calculated using the formula below.

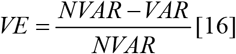

Where VE is the Vaccine Effectiveness, NVAR is the Attack rate among the unvaccinated, and VAR is the Attack rate among the vaccinated.

### Ethical Considerations

The study was approved by the College of Medicine Research Ethics Committee (COMREC) under the protocol number of P.O7/24-0923.

## Results

### Frequency of Cholera cases

Table 1 presents the background characteristics of the cholera cases, including sex, age group, vaccination status, disease outcome, dehydration level and occupation. A total of 28,920 cholera incidents, all of which had established vaccination histories and disease outcome statuses, were analysed.

**Table 1:** Frequency of Sex, Age Group, Vaccination and Disease outcome among suspected cholera cases.

There were 55.89% males and 43.87% females, with 72.10% aged 15 years or older; 98.04% were unvaccinated, compared to 1.96% who had received vaccination. 47% were severely dehydrated, 97.27% recovered (alive), and 2.73% were documented as deceased(dead), yielding a cholera case fatality rate (CFR) of 2.73%.

### Distribution of cholera suspected cases by district

#### Distribution of suspected cholera cases by District

Figure 1 presents the distribution of suspected cholera cases by district. The predominant proportion of cases with ascertainable vaccination status originated from Blantyre (26%), followed by Salima (12%), and the least were Mzimba South and Likoma, 0.01% each

**Figure 1.**
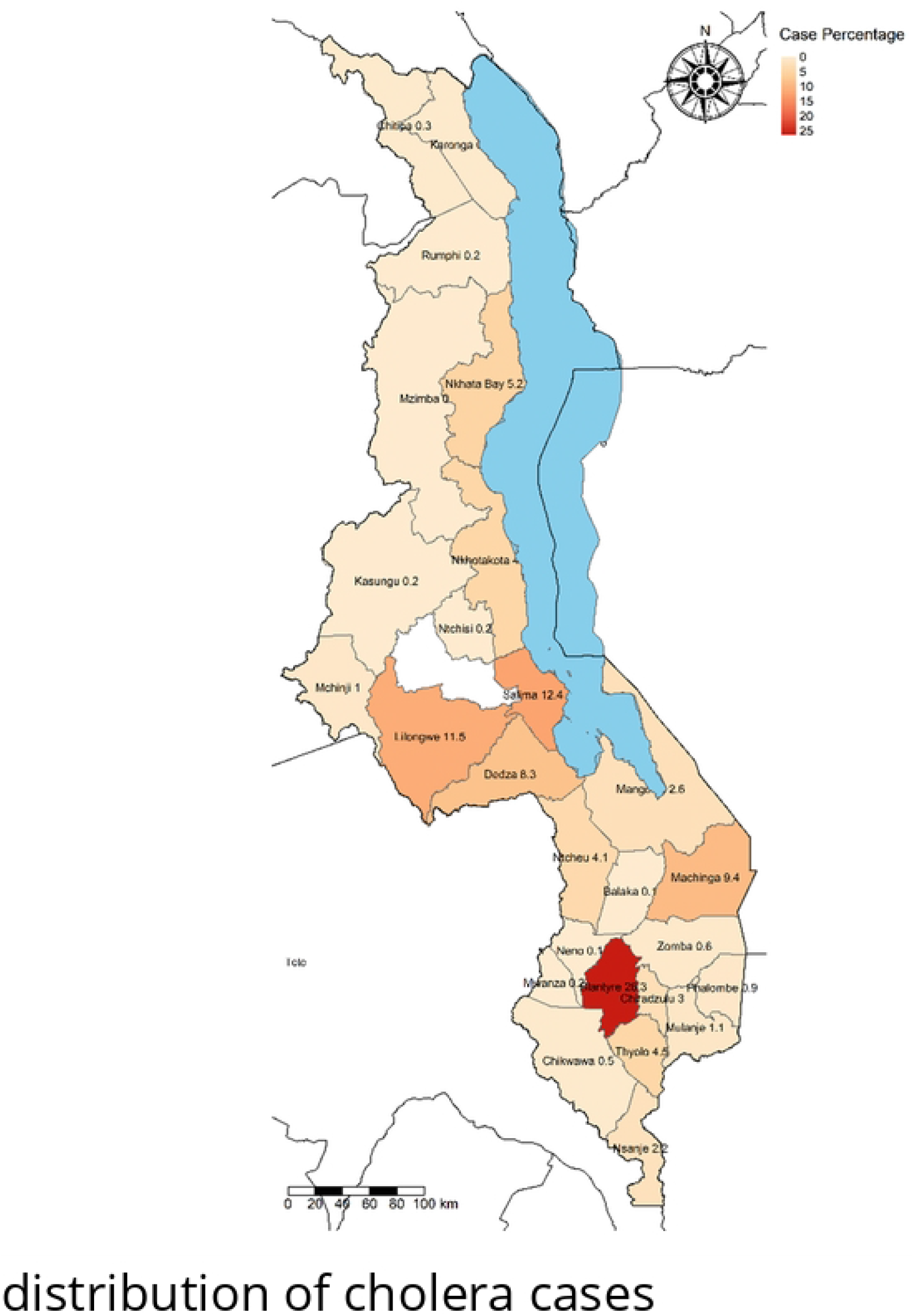
Distribution of suspected cholera cases

**Figure 2.**
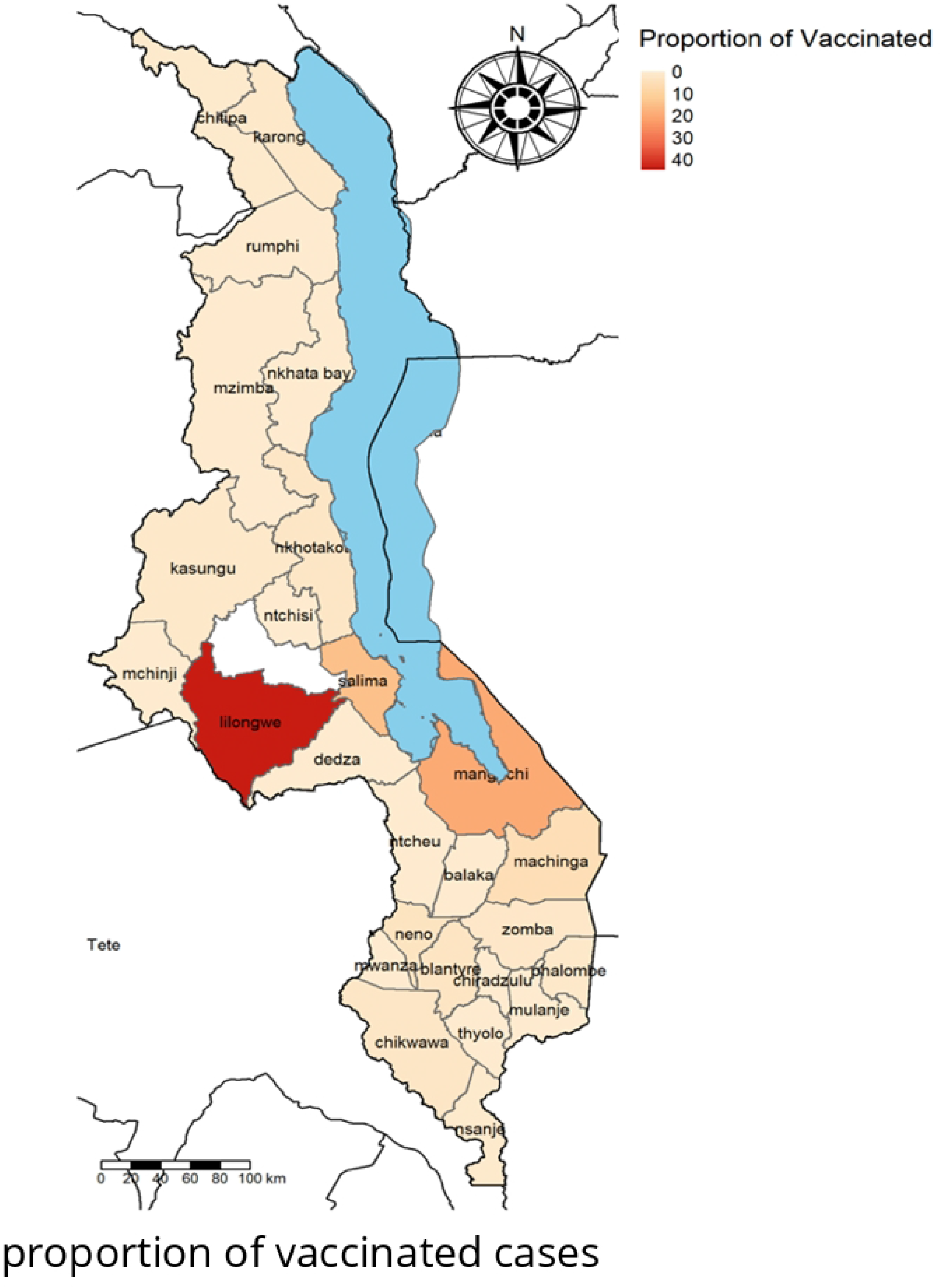
Percentage of Vaccinated Individuals per District

### Cross-tabulation of suspected cholera cases by demographic variables against vaccination status

**Table 2** shows the distribution of demographic and clinical characteristics by vaccination status. Most cases were unvaccinated. Vaccinated cases were more frequently female and younger, with a higher proportion in the 0–4 and 5–14 age groups compared to unvaccinated cases (p < 0.001). The proportion of deaths was lower among vaccinated cases than among unvaccinated cases (1.2% vs 2.8%; p=0.038). Severe dehydration was less common among vaccinated individuals(p<0.001). Occupational distributions differed significantly by vaccination status; however, occupation was unknown for most cases (p<0.001).

**Table 2:** Oral Cholera Vaccination coverage among suspected cholera patients.

### Cross-tabulation of cholera suspected cases by district against vaccination status

Table 3 shows OCV coverage per district. The OCV coverage in Liloma and Neno was estimated at 100%. This was due to low records from the two districts. Mangochi 15.25%, while Balaka, Chiradzulu, Dedza, Mchinji Mulanje, Mwanza, Mzimb South, Ntcheu, Ntchisi and Thyolo had 0%

**Table 3:** Oral Cholera Vaccination Coverage per District.

### Proportion of Vaccination Status per District

Figure 5 shows the proportion of the 568 vaccinated cholera cases. 44.37% were from Lilongwe, followed by Mangochi (19.89%). Balaka, Chiradzulu, Dedza, Mchinji, Mulanje, Mwanza, Mzimba South, Ntcheu, Nthchisi, and Thyolo each had zero vaccinated cases.

### OCV effectiveness across districts

Table 4 shows the effectiveness of OCV across the districts. The overall effectiveness was calculated to be 98.06%. High effectiveness was observed in Thyolo, Nkhata Bay, Mwanza, Mulanje, Mchinji, Chiradzulu and Balaka, each having 100%. The lowest was 81.25% in Mangochi.

**Table 4:** Table 4 OCV effectiveness across districts.

### Cross-tabulation of cholera suspected cases by demographic variables against levels of dehydration

Table 5 presents dehydration levels among 21,869 cholera cases; 11,438 (52.3%) had moderate and 10,431 (47.7%) had severe dehydration. Sex and Occupation were not associated with dehydration severity (p> 0.9). Age group was significantly associated with severity (p < 0.001), with a higher proportion of severe cases among individuals aged ≥15 years. Mortality was higher among severe cases (2.5%) compared with moderate cases (1.8%; p < 0.001). Vaccinated individuals were less frequently observed among severe cases (0.9%) than among moderate cases (1.7%; p < 0.001).

**Table 5:** Levels of dehydration of Suspected Cholera Cases.

### Cross-tabulation of cholera suspected cases by district against dehydration levels

Table 6 presents the distribution of cholera cases across districts by dehydration level, which varied substantially by district. Moderate cases were predominantly reported from Salima (31%), Blantyre (26%), and Dedza (18%), while severe cases were concentrated in Blantyre (44%), Thyolo (12%), Chiradzulu (7.5%), Nkhotakota (6.7%), and Nkhata Bay (8.1%). Several districts reported cases in only one severity category, with Dedza and Salima reporting no severe cases, and Balaka, Chikwawa, and Chitipa reporting no moderate cases

**Table 6:** dehydration levels of suspected cholera cases per District.

### Multivariable analysis of levels of dehydration against Sex, Vaccination, Age group, and Occupation

Table 7 presents crude analyses, showing that death was associated with higher odds of severe dehydration (OR = 1.39; 95% CI: 1.16–1.68); however, this association was not retained after adjustment (adjusted OR = 0.72; 95% CI: 0.45–1.12). Vaccination was associated with lower odds of severe dehydration in the crude model (OR = 0.50; 95% CI: 0.39–0.64), but this association did not remain significant after adjustment. Sex was not associated with dehydration severity in either crude or adjusted analyses. Compared with children aged 0–4 years, individuals aged 15 years and older had increased odds of severe dehydration in the crude analysis (OR = 1.12; 95% CI: 1.02–1.23), though age group was not associated with dehydration severity after adjustment. In contrast, occupation remained associated with dehydration severity, with farmers showing higher odds of severe dehydration in both crude (OR = 2.08; 95% CI: 1.45–3.01) and adjusted analyses (adjusted OR = 2.02; 95% CI: 1.41– 2.94). Casual workers and fishermen had lower odds of severe dehydration in both models, while estimates for several occupational categories were unstable due to sparse data

**Table 7:** Estimates of Crude and Adjusted ORs by Sex, Age Group and Occupation against dehydration levels.

The vaccine effectiveness towards reducing severe cholera among suspected cholera cases was 50% (95% CI: 36-61)

## Discussion

Oral cholera vaccine is one of the strategies to complement WASH, Surveillance, and IPC intervention. Due to the global shortage of OCV doses, the ICG recommended the temporary use of one dose of OCV. Malawi conducted an OCV campaign during the 2022-2023 cholera outbreak in a reactive model. A national OCV survey report of 2023 estimated OCV coverage at 55.40% among the community members, of whom 82.20% received a single dose [18]. In this study, OCV coverage among cholera suspects was at 1.97%, a close reflection of what Grandesso et al found among cases in a case-control study conducted at Lake Chikwa, which estimated the coverage among cases to be 1.80%[17]. This indicates that there was low OCV coverage among the cholera suspects captured at the health facility during the 2022-2023 cholera outbreak, with 98.04% unvaccinated.

A single dose of OCV Euvichol-plus showed vaccine effectiveness (VE) of 98% in reducing cholera incidence and vaccine effectiveness of 50%(36-61, p<0.001) towards reducing severe cholera cases reported at health facilities during the 2022-2023 cholera outbreak. It showed that using a single dose of OCV Euvichol-plus during an outbreak is of advantage, and it complements other interventions. Our finding was aligned with the 52.7% single-dose VE of Euvichol-plus that was estimated in the DRC [23]. It presumes a similarity with a two-dose vaccine effectiveness towards reducing severe dehydration among attended cholera cases, which was found at 62% protection against cholera infection across all ages and 79% among individuals above 5 years in Bangladesh[13]. While in Zambia, the VE was estimated at 81% for the age group one year and above[24]. All these evaluations were done on a reactive OCV campaign towards the cholera outbreak. The studies done in Bangladesh and Zambia used a two-dose strategy during the cholera outbreak. All the studies show that OCV helps in reducing Cholera incidence during outbreaks. And the use of a single dose can be of advantage when there is a shortage of OCV at the global level.

Sex disparity is prevalent in disease intervention within the community. Mostly, this is due to differences in disease risk perception between females and males[22]. This difference affects the uptake of interventions. We found that females had a higher OCV coverage of 2.30% compared to 1.7% in males. Females were 1.26(95%CI: 1.13;1.38) likely to be vaccinated compared to males. The post-OCV coverage survey also found that most individuals who received the OCV were female (57.90%)[18].

## Study limitations

Our study was prone to recall bias, which we could not control. Vaccination status in the cholera line list was documented based on the patient’s memory. Data clerks were not confirming the vaccination status in the line list with the patient’s OCV vaccination card.

During a cholera outbreak, testing to confirm a cholera positive through culture is not done on every suspected cholera case. Hence, the likelihood of including false positive cholera cases in the cholera line list was very high. False positive cholera cases would arise due to variation in the definition of confirmation. The definition of confirmation changed during the outbreak period, where three confirmation criteria were used: culture test, Rapid Diagnostic Test (RDT), and epidemiological link. The use of epidemiological links is subjective.

## Conclusion

We used a national cholera line list to estimate OCV coverage among cholera cases presented at different health facilities in Malawi during the 2022-2023 cholera outbreak. 28,920 records of suspected cholera cases with known vaccination status and disease outcome were included to estimate OCV coverage among cholera cases.

Overall OCV coverage among suspected cholera cases during the 2022-2023 cholera outbreak was low (1.97%). This indicated that the majority of cholera suspected individual reported at health facilities during the 2022-2023 cholera outbreak were not vaccinated. Among the vaccinated cholera suspects, the majority were moderate; the severe cases showed that they were not vaccinated.

The results of this study showed that one dose of OCV has a protective factor against cholera infection and severity. In this study, the VE was 98%, quite high and in agreement with other studies done evaluating the effectiveness of the Euvichol-plus vaccine towards cholera outbreaks. The vaccine effectiveness was found at 50% towards reducing severe dehydration cholera cases. However, a gap still exists in knowledge on the duration of Euvichol-plus single-dose vaccine protection against cholera infection.

## Data Availability

All the data used for the analysis will be provided.

## Acknowledgement

We would like to acknowledge the Public Health Institute of Malawi (PHIM) and the Expanded Program on Immunisation for the support rendered during the development of this manuscript

## Notes

### Competing Interest Statement

The authors have declared no competing interest.

### Funding Statement

we used secondary data, and there was no funding towards this work.

